# ANMerge: A comprehensive and accessible Alzheimer’s disease patient-level dataset

**DOI:** 10.1101/2020.08.04.20168229

**Authors:** Colin Birkenbihl, Sarah Westwood, Liu Shi, Alejo Nevado-Holgado, Eric Westman, Simon Lovestone, Martin Hofmann-Apitius, on behalf of the AddNeuroMed Consortium

## Abstract

**Background:** Accessible datasets are of fundamental importance to the advancement of Alzheimer’s disease (AD) research. The AddNeuroMed consortium conducted a longitudinal observational cohort study with the aim to discover AD biomarkers. During this study, a broad selection of data modalities was measured including clinical assessments, magnetic resonance imaging, genotyping, transcriptomic profiling and blood plasma proteomics. Some of the collected data were shared with third-party researchers. However, this data was incomplete, erroneous and lacking in interoperability.

**Methods:** We systematically addressed several limitations of the originally shared data and provide additional unreleased data to enhance the patient-level dataset.

**Results:** In this work, we publish and describe ANMerge, a new version of the AddNeuroMed dataset. ANMerge includes multimodal data from 1702 study participants and is accessible to the research community via a centralized portal.

**Conclusions:** ANMerge is an information rich patient-level data resource that can serve as a discovery and validation cohort for data-driven AD research, such as for example machine learning and artificial intelligence approaches.

ANMerge can be downloaded here: https://doi.org/10.7303/syn22252881

## 1. Background

Alzheimer’s disease (AD) is a progressive disease whose pathology develops years before cognitive symptoms arise and a diagnosis is made by a clinician (Sperling et al., 2011). Early intervention in non-cognitively impaired, pre-symptomatic disease stages is instrumental to any future disease modifying therapy. Enabling such an early intervention poses the problem of diagnosing a patient with AD before cognitive symptoms indicate disease presence. One approach to establish whether a specific individual is in the pre-symptomatic stages of the disease is a diagnosis based on informative disease biomarkers. The critical prerequisite for discovery and validation of such biomarkers are resourceful patient-level datasets (Morgan et al., 2019). However, findable AD cohort datasets which are accessible to the research community are scarce.

Open science is a paradigm aimed at increasing societal benefit of research through dissemination and sharing of scientific data. This enables usage and analysis of collected data by the whole research community which subsequently will increase the achieved knowledge gain. Currently, the prime example of following the open science paradigm in the AD field is the Alzheimer’s Disease Neuroimaging Initiative (ADNI; Mueller et al., 2005).

ADNI is an information rich, comprehensive clinical AD cohort dataset that enables secure, yet easy access to its patient level data for researchers with reasonable study interest. In only a few days raw data as well as a preprocessed version of ADNI (ADNIMERGE) are accessible via the Laboratory of Neuro Imaging (LONI) service (http://loni.usc.edu/). Initial preprocessing, arranging and cleaning of data is the most time consuming step in data analysis. Due to that, a major cumulative time save is possible by sharing an already preprocessed, easy to analyse dataset instead of a raw data collection. Here, researchers can simply use the provided ADNIMERGE and thereby avoid investing additional time into data preprocessing and cleaning.

While ADNI is a tremendously important resource, as every cohort dataset it comes with its own limitations and biases (Whitwell et al., 2012, Ferreira et al., 2017). To ensure reliability of observations made in one cohort, validation in data from independent cohorts is necessary (Fröhlich et al., 2018). Still, apart from ADNI there are not many AD cohort studies which 1) share their data in a similarly comprehensive version and 2) keep the bureaucracy during an access application as straightforward as ADNI does. From our experience, access applications are often time consuming and if access is granted, shared data is sometimes lacking important information. Therefore, other easily accessible and information rich alternatives besides ADNI are crucial.

In 2005, Lovestone et al. started AddNeuroMed, an Innovative Medicine Initiative (IMI) funded project which involved gathering longitudinal patient data at multiple sites across Europe (Lovestone *et al*., 2007). Its main goal was to identify urgently needed progression biomarkers for AD. For this purpose, a broad spectrum of variables was measured including demographics, neuropsychological assessments, genetic variations and transcriptomics, blood plasma proteomics, and structural magnetic resonance imaging (MRI) of the brain. In 2015 a subset of the collected data was uploaded on Synapse (https://www.synapse.org/). Next to the original AddNeuroMed data, some data from participants of the Maudsley BRC Dementia Case Registry at King’s Health Partners cohort (DCR) and the Alzheimer’s Research Trust UK cohort (ART) was included (Hye *et. al*., 2006). Although the shared AddNeuroMed collection is a large dataset, involving more than 1700 participants, it has only been cited 64 times^1^. In contrast, ADNI, which involves roughly 2400 individuals, was cited more than 1300 times^2^. Compared to the impact ADNI has had on recent research activities, it seems AddNeuroMed does not reach its full potential. One probable reason for the comparably lower data usage might be the findability and the state of the data published on Synapse. The dataset 1) has never been officially published, 2) is not easy to work with due to missing organisation, and 3) is not complete with several entries being erroneous or lacking information. To enable the research community to leverage the full potential of this dataset, a lot of data preprocessing efforts are needed and it is vital to point the community towards this unsalvaged resource.

In this work, we present and publish a new, improved and updated version of AddNeuroMed called ANMerge. ANMerge is a comprehensive, preprocessed AD cohort dataset which is again accessible via Synapse (https://www.synapse.org/#!Synapse:syn4907804). It is fully interoperable in between its modalities and rigorous data curation was performed to ensure higher information density and usability. Furthermore, we present a detailed overview on which and how much data is available in the dataset. Finally, we highlight the increased preprocessing efforts involved in creating such a dataset. By making ANMerge accessible, we aim to provide the AD research community with an information rich alternative to previously published cohort datasets, and thereby support the discovery and robust validation of scientific insights.

## 2. Methods

### 2.1 Data collection

AddNeuroMed data collection was performed at six different centers across Europe: University of Kuopio, Finland; Aristotle University of Thessaloniki, Greece; King’s College London, United Kingdom; University of Lodz, Poland; University of Perugia, Italy; and University of Toulouse, France (Lovestone et al., 2007). The participation of those centers highlights AddNeuroMed as a major cross-european effort in AD related data collection. At each site, all protocols and procedures were approved by Institutional Review Boards and informed consent was obtained for all patients according to the Declaration of Helsinki (1991) (Simmons et al., 2009). In cases where dementia compromised capacity assent from the patient and consent from a relative, according to local law, was obtained.

Exclusion criteria included other neurological or psychiatric diseases, significant unstable systemic illness or organ failure, and alcohol or substance misuse. AD diagnosis followed the Diagnostic and Statistical Manual for Mental Diagnosis, fourth edition and National Institute of Neurological and Communicative Disorders and Stroke–Alzheimer’s Disease and Related Disorders Association criteria (McKhann et al., 1984). AD patients were included if they exhibited a Mini Mental State Examination (MMSE) score in the range of 12-28, a Clinical Dementia Rating (CDR) scale score of above 0.5, and were aged 65 years or above. Individuals were considered as MCI according to the Petersen criteria (Petersen, 2004). For inclusion, MCI patients aged 65 or above, the MMSE score ranged between 24 and 30, and they scored 0.5 on the CDR. Participants were considered to be cognitively healthy if they showed normal performance on cognitive tests (within 1,5 SD of average for age, gender and education) and scored 0 on the CDR (Hye et al., 2014).

AddNeuroMed’s study protocols were designed to be at least partially compatible to ADNI (Lovestone et al., 2007). **Figure 1** illustrates when data collection was performed for each modality.

**Figure 1:**
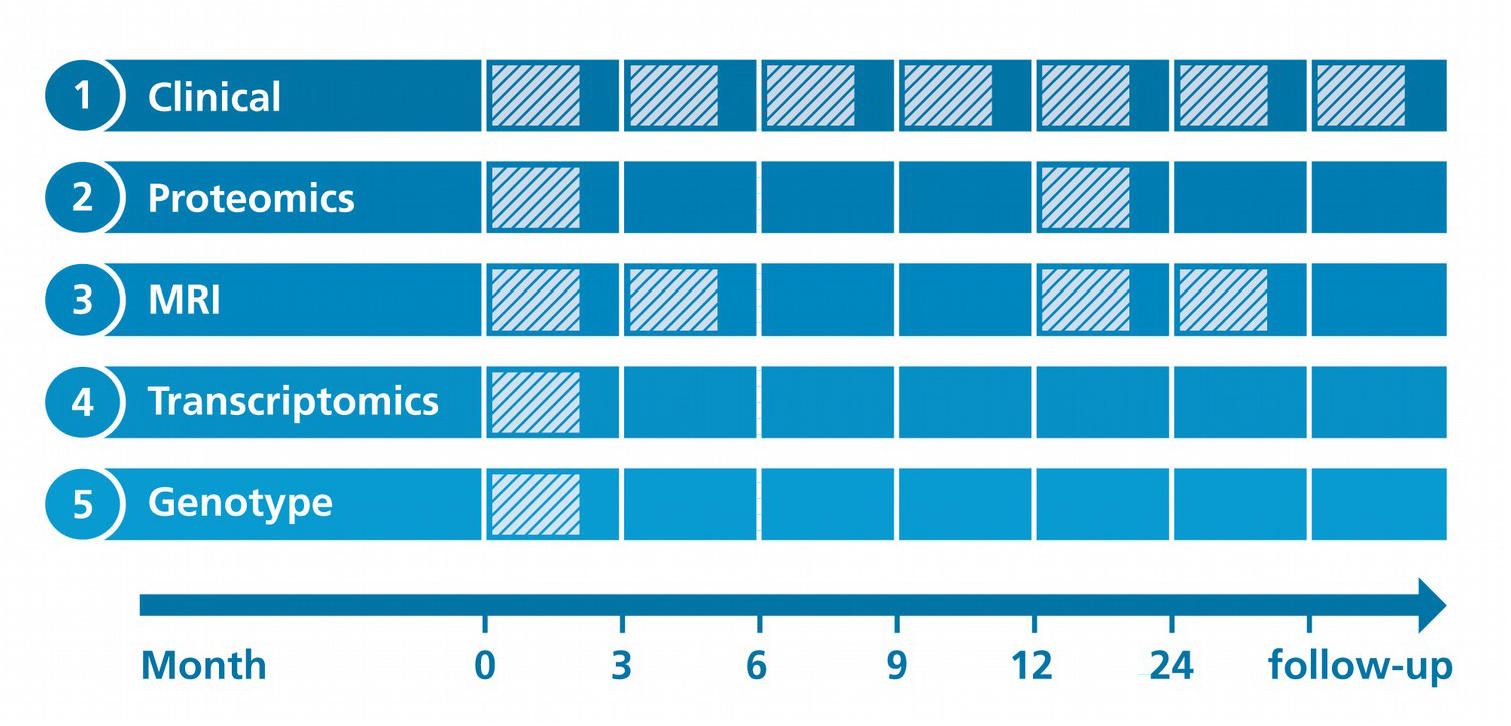
Overview on longitudinal data collection per modality. **Proteomics:** Proteomic data from blood plasma,. **Transcriptomics:** Transcriptomic data from blood plasma.

#### Clinical assessments

At each participants visit throughout the study, a broad collection of neurocognitive and psychological assessments were performed, including the MMSE, CDR, GDS (Geriatric Depression Scale), NPI (Neuropsychiatric Inventory), ADAS-Cog (Alzheimer’s Disease Assessment Scale-Cognitive Subscale), ADCS-ADL (Alzheimer’s Disease Cooperative Study Activities of Daily Living Scale), the full CERAD battery (Morris et al., 1989), the Hachinski Ischemic Score and the Webster Rating Scale. The frequency with which assessments were made varied between diagnostic groups. During the first year, AD cases completed assessments every three months and annual follow-up visits afterwards. MCI patients and healthy individuals from AddNeuroMed, as well as all participants from the ART and DCR cohorts were assessed regularly every twelve months.

#### Proteomics

Proteomic data were measured in blood plasma using a Slow Off-rate Modified Aptamer (SOMAmer)-based array called ‘SOMAscan’ (SomaLogic, Inc, Boulder, Colorado). Data collection was performed at baseline and again one year into the study. Details on data acquisition are presented in Kiddle et al. (2010) and Sattlecker et al. (2014). In brief, using chemically altered nucleotides the protein signal is turned into a nucleotide signal that can be measured using microarrays. Per sample 8 µL plasma were required and levels of 1001 distinct proteins were measured. An in-depth description of the array technology can be found in Gold et al. (2010).

#### Genotyping

AddNeuroMed participants were genotyped in two batches. For batch one the Illumina HumanHap610-Quad Beadchip was used, while batch two was processed using the Illumina HumanOmniExpress-12 v1.0. More information can be found in the method section of Loudursamy et al. (2012) and Proitsi et al. (2014). All genotyping was performed at the Centre National de Génotypage in France.

#### Transcriptomics

Blood samples for the collection of gene expression data were taken at study baseline. Transcriptional profiling was performed in two batches using the Illumina HumanHT-12 v3 (batch one) and v4 (batch two) Expression BeadChip kits. Original raw data can be found in GEO^3^. Preprocessed raw data files, as well as post quality control, batch corrected expression values, are distributed via Synapse. The processed data underwent background correction, log base two transformation and all values were robust spline normalized (Voyle et al., 2016). Outlying samples were excluded. Batch correction was performed using ComBat (Johnson et al., 2007). All data were subset to probes that could reliably be detected in at least 80% of samples in at least one diagnostic group. More details on the processing of the data is explained in Voyle et al. (2016).

#### Magnetic Resonance Imaging

1.5 Tesla T1-weighted MRI images were taken at three different timepoints throughout the study (Month 0, 3, 12). The first 3 month interval was explicitly chosen to contrast the 6 month MRI follow-up of ADNI and thereby evaluate if 3 month could potentially be enough to observe substantial changes in brain structure. Protocols for imaging were aligned to the ADNI study. Details on the AddNeuroMed MRI data acquisition have been described in Simmons et al. (2011 and 2009). ANMerge provides access to collected raw images as well as processed brain volumes and cortical thickness calculated using FreeSurfer 5.3 and 6.0.

### 2.2 Data preprocessing

First, we manually investigated all raw data files and evaluated the availability and state of all data types. Manual changes to the data were avoided by performing all data alternating steps computationally.

We tried to build the most informative and complete, yet minimally complex, version of AddNeuroMed possible. Therefore, we carefully selected features from the raw data for inclusion into the new ANMerge version. To reduce the number of variables in ANMerge, we only included total scores of clinical assessments instead of listing all sub-scores and individual answers. Variables not considered in the new dataset are still accessible through Synapse.

Not all participants from the DCR and ART cohorts underwent data collection in the course of AddNeuroMed. However, since clinical assessments between the original AddNeuroMed study and DCR were largely overlapping, we decided to include all DCR participants into ANMerge, even if they lacked other modalities apart from clinical data. From the ART cohort, we only included individuals who had been assessed in at least one modality next to clinical data to reduce data sparsity.

The new ANMerge dataset is divided into modality specific subtables which makes unimodal analysis straightforward. To enable full interoperability between these subtables, we mapped all previously modality specific participant identifier to a single common one. Additionally, we enriched ANMerge with data previously not available in the Synapse version. For example, we added missing diagnoses and clinical assessment scores as well as months in study as an unambiguous time scale. Previously, only visit numbers were reported. The latter are misleading due to differences in assessment intervals between diagnostic groups (e.g. visit 2 for healthy and MCI participants corresponds to visit 5 of AD patients). Information which is no subject to change (e.g. APOE genotype) was added to all entries of a participant in order to reduce sparsity. Furthermore, to increase interoperability not only within AddNeuroMed itself but also to other data resources, we mapped variable names to public database identifiers wherever possible. For example, proteomic variables are given as UniProt identifiers, genotype data is encoded as rs-numbers and gene expression probes as Illumina IDs (Du et al., 2008). All of these identifiers can be easily mapped to other resources and be enriched with information from public databases.

## 3. Results

### 3.1 Overview on Data

The resulting ANMerge dataset comprises four data modality specific subtables, genotype data in PLINK format and one combined table providing all preprocessed information as one. Respectively, one subtable was created for clinical data, proteomics, FreeSurfer calculated MRI features, and gene expression values. Next to diagnosis and clinical assessments, the clinical subtable also provides participants demographics, family history and medication data.

In total the dataset comprises information on 1702 patients, out of which 773, 665 and 264 originated from the AddNeuroMed, DCR and ART cohorts respectively (Table 1). Data on 4585 individual participant visits are reported. At study baseline, 512 participants had been diagnosed with AD, 397 with MCI and 793 were non cognitively impaired individuals. Table 1 describes the average characteristics of each diagnosis group at baseline. On average, cognitively affected individuals (i.e. MCI and AD) in ANMerge were 77 years old at baseline, completed 9.7 years of full-time education and 59% of them were female. Healthy individuals averaged to an age of 74.5 years, underwent 12.3 years of education and 59% are female. During study runtime 48 and 11 healthy participants converted to MCI and AD respectively. Out of all patients diagnosed with MCI at baseline 70 converted to AD.

**Table 1:** Summary statistics describing the ANMerge cohort at baseline. **N:** Number of participants with corresponding diagnosis. **ANM:** Number of participants originally from the AddNeuroMed study, **DCR:** Number of participants originally from the DCR study. **ART:** Number of participants originally from ART study. **CTL:** Healthy control participants

| Diagnosis | N | ANM | DCR | ART | Age (SD) | Female % | Education (SD) | APOE e4 positive % |
| --- | --- | --- | --- | --- | --- | --- | --- | --- |
| CTL | 793 | 266 | 423 | 104 | 74.5 (6.4) | 59 | 12.3 (4.3) | 25 |
| MCI | 397 | 247 | 89 | 61 | 76.0 (6.5) | 55 | 10.0 (4.3) | 40 |
| AD | 512 | 260 | 153 | 99 | 78.6 (7.2) | 63 | 9.4 (4.3) | 54 |
| Total | 1702 | 773 | 665 | 264 | 76.4 (6.9) | 59 | 10.9 (4.5) | 39 |

Not every study participant took part in data collection of all modalities. For our evaluation, we considered participants as represented in a modality if at least one modality specific variable was measured. This implies that not necessary all variables of that modality were available for a given participant (e.g. an individual listed in the clinical table might have MMSE scores but no ADAS-Cog). We found that clinical data is reported for all 1702 participants, while MRI, proteomic, gene expression, and genotype data were collected for subsets of several hundred participants each (Table 2 ‘Subjects’). Figure 2 demonstrates the number of patients assessed across multiple modalities. In total, 235 participants have been assessed with regard to all five data modalities. By reducing the number of modalities included into an analysis, subsequently the number of available participants rises. For example, when conducting a multimodal study using transcriptomic, genotype and clinical variables data from 484 participants would be available. Focusing only on genotype and clinical data yields 640 analyzable subjects.

**Table 2:** Content of modality subtables.

| Modality | Subjects | Features |
| --- | --- | --- |
| Clinical | 1702 | 40 |
| Proteomics | 680 | 1016 |
| MRI | 453 | 136 |
| Gene expression | 709 | 56701 |
| Genotype | 644 | 789470 |

**Figure 2:**
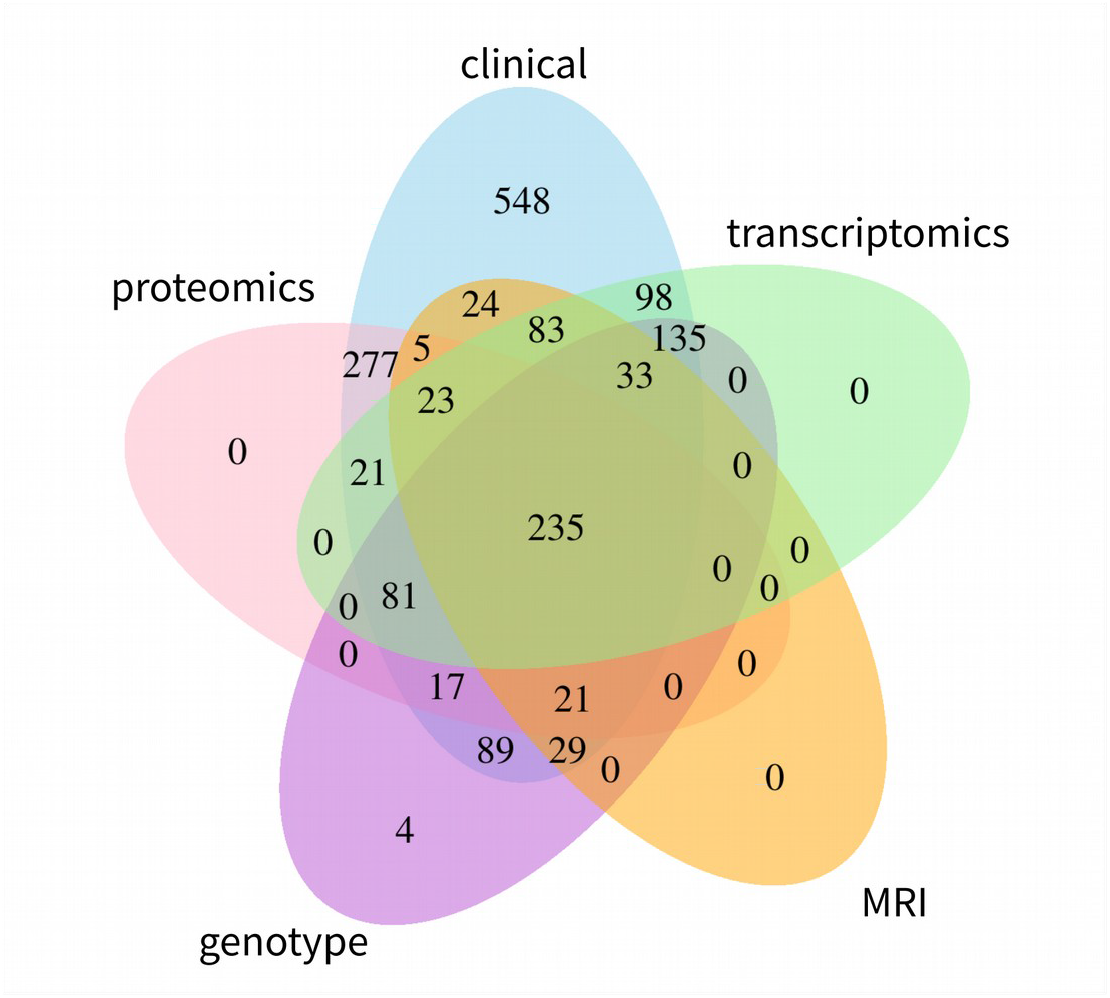
Participant overlap across modalities. The numbers illustrate the amount of participants with available information.

All in all, data on more than 800,000 variables are reported in ANMerge. 40 of them correspond to the clinical modality, 56701 originate from gene expression analysis, 136 are MRI variables, and 1016 were assessed in blood proteomics (Table 2 ‘Features’).

As most clinical studies, AddNeuroMed exhibits a declining number of participants over study runtime (Figure 3). For most patients (n = 1136) at least one additional visit 12 months after baseline is available in the data. The drop of AD patients at month 3 to 9 is explained by the fact that only AD cases recruited in the original AddNeuroMed study had three monthly visits during the first year, while ART and DCR assessed all patients annually. The longest follow-up exhibited in the data spanned 12 years.

**Figure 3:**
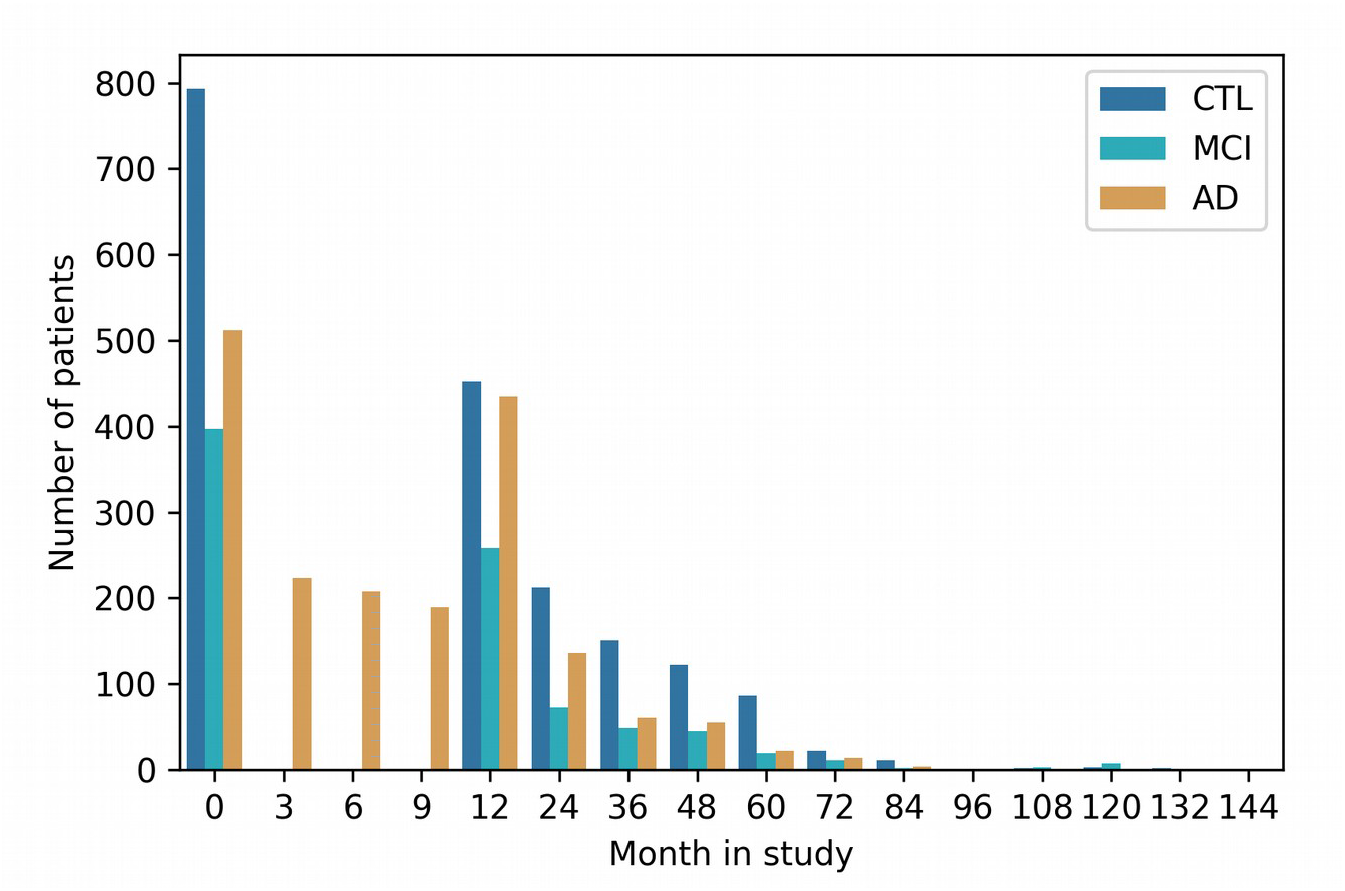
Longitudinal follow-up and patient drop-out throughout study runtime per diagnosis group. **CTL:** Healthy controls. **MCI:** Mild cognitive impaired participants. **AD:** Alzheimer’s disease patients.

### 3.2 Data After Preprocessing

During the preprocessing and curation of AddNeuroMed we addressed multiple issues detected in the previous version. The previous version of AddNeuroMed was indexed using distinct patient identifier across its modalities, thereby impeding multimodal analysis due to missing internal interoperability. Standard data integration techniques like table joins were impossible. By mapping all present identifiers to a unique one, we enabled inter-modality interoperability such that tables can now easily be analyzed together. Additionally, we provide a new identifier mapping file which helps to map the unified identifiers to the raw data for backwards compatibility. The exact visit dates and time scale (months in study) we added to patient entries make longitudinal follow-up easier to understand and time differences less misleading than relying on previously reported visit enumeration. Information that will stay permanent (e.g. APOE e4 status) throughout study runtime is now reported at every visit for that respective patient, not only at baseline. Multiple issues found in the data (e.g. typos and erroneous entries) have been corrected.

Although for example proteomic and transcriptomic data were presented for some DCR and ART participants in the previous AddNeuroMed version, no corresponding clinical data was available, including important information like participant diagnosis. ANMerge now has all available clinical data for the two associated cohorts, which critically increases the amount of actionable information in the dataset.

### 3.3 Accessing ANMerge

ANMerge and the underlying data are available under https://www.synapse.org/#! Synapse:syn4907804. To ensure data privacy a straight-forward data access application has to be completed. Access approval takes roughly seven days. These data also provide subscores and individual answers for each clinical assessment / questionnaire.

## 4. Discussion

In this work, we presented ANMerge, a longitudinal multimodal AD cohort dataset that we made accessible to the research community. Since the most time consuming part about data analysis is often the preprocessing of data, we believe that the cumulative time save, achieved by sharing readily preprocessed datasets, can lead to faster global scientific advancement. Additionally, by describing the characteristics of the available our dataset in detail, we aim to enable researchers to evaluate on first sight if ANMerge is suited for their analysis.

Establishing reliable results through external validation on independent cohorts is of utmost importance, especially when dealing with high complex diseases like AD. Up to date, and to the best of our knowledge, the vast majority of data-driven approaches in AD rely solely on ADNI data. To validate discoveries made in ADNI on other datasets, a high overlap in measured variables is a prerequisite. Previously, we could demonstrate that despite evident differences to ADNI, ANMerge is a viable validation dataset (Birkenbihl *et al*., 2020).

Recently, more studies such as PREVENT-AD (Tremblay-Mercier et al., 2014) and EPAD (Solomon et al., 2018) joined the ranks of ADNI, DIAN (Morris et al., 2012) and others by making their data accessible to third party researchers. The currently running Deep Frequent Phenotype Study (Lawson et al., 2017) already emphasized that collected data will be published. This shift in the AD data landscape towards increasingly accessible datasets is an important development for the goal of finding reliable biomarkers.

## 5. Limitations

While AddNeuroMed is undoubtedly a valuable dataset, it still has some noteworthy limitations. The main limitation of the data is that the amyloid statues of participants is unknown. No positron emission tomography (PET) imaging was performed and cerebrospinal fluid markers were not assessed.

As in many clinical cohort datasets, missing data is a considerable issue in AddNeuroMed. Not every patient was involved in the assessment of every data modality and within a modality not necessarily all variables were measured for each patient.

Compared to ADNI, AddNeuroMed lacks comprehensive documentation. Retrospectively searching for study procedures and protocols of an already concluded, older cohort study proved to be very difficult. The original study website is not available anymore and exhaustive study protocols were not findable. However, we tried to address this limitation by collecting and assembling all available information and links in this publication. While the original AddNeuroMed dataset provided descriptive data dictionaries for most clinical variables, we extent the documentation by meaningful connections of other modalities to public databases (e.g. UniProt or dbSNP) by mapping their variable names to appropriate identifiers wherever possible.

The genotype and transcriptomic data presented in ANMerge was acquired in two separate batches of participants. This implies that the data can be subject to systematic batch effects and appropriate adjustments should be made (Benito et al., 2004).

## 6. Conclusion

By publishing ANMerge, we want to follow the open science paradigm and contribute to a culture of data sharing in AD research. Participation in observational clinical cohort studies is a huge investment by volunteering patients and healthy individuals. They undergo extensive and sometimes intrusive repeated measurements, most of the times without any direct benefit for the individuals themselves, with the ultimate aim to contribute to disease research. We believe that it is an ethical imperative to honor their investment by enabling that their data is used to generate the most societal benefit possible.

## Data Availability

All data are available under: https://doi.org/10.7303/syn22252881

## Declarations

### Ethics approval and consent to participate

All data collection was performed in following the Declaration of Helsinki and informed consent was gathered.

## Consent for publication

Data were already published previous to this work and participants agreed to publication to generate the most societal benefit.

## Availability of data and materials

All data are available under: https://doi.org/10.7303/syn22252881

## Authors contributions

Drafted manuscript: CB

Research design: CB, MHA, SL

Data processing: CB, EW, SW, LS

Data collection: EW, SL

Revised manuscript: SW, LS, ANH, EW, MHA

## Acknowledgements

The authors of this article wanted to thank every participant and professional involved in AddNeuroMed, DCR and ART data collection.

## Footnotes

1 18.09.2019 PubMed query: “AddNeuroMed” AND (“Alzheimer’s disease” OR “dementia”); state

2 18.09.2019 PubMed query: “ADNI” AND (“Alzheimer’s disease” OR “dementia”); state

3 Batch one: https://www.ncbi.nlm.nih.gov/geo/query/acc.cgi?acc=GSE63060 Batch two: https://www.ncbi.nlm.nih.gov/geo/query/acc.cgi?acc=GSE63061

